# Intravital mid-infrared biosensing by normalized spatial probing of self-referenced optothermal signals

**DOI:** 10.64898/2026.05.27.26354202

**Authors:** Constantin Berger, Björn Puttfarcken, Jiawang Qiu, Isabella Hauer, Sebastian Herr, Dominik Jüstel, Miguel A. Pleitez

**Author notes:** equal contributions (Constantin Berger, Björn Puttfarcken).

## Abstract

We present a compact pump-and-probe mid-infrared Optothermal Spectrometer (OTHES) equipped with Spatial Probing and Autocorrection (SPAC) optimized for robust intravital application in humans. SPAC-OTHES facilitates alignment stability and spectral comparability across different measurement sessions involving different skin types. Contrary to state-of-the-art, SPAC-OTHES uses camera-based beam detection and an auto-calibration mechanism that enables ca. 73% better spectral reproducibility in intravital measurements in human volunteers than non-calibrated readouts. Moreover, SPAC-OTHES has the potential to lower the glucose quantification error, as demonstrated here in artificial skin phantoms, where an improvement of 52% compared to conventional diode-based detection was observed. The compactness of OTHES, combined with reliable SPAC-readout, has the potential to accelerate commercialization and broad application of biosensors based on mid-infrared spectroscopy.

## Introduction

Vibrational spectroscopy by mid-infrared (mid-IR) excitation enables non-destructive assessment of molecular composition in biological tissues without the need for exogenous labels, using instead the distinct spectral fingerprints of biomolecules (1). Benefiting from its speed and robustness, mid-IR spectroscopy has been widely applied for use-cases including industrial quality control (e.g., chemical and pharmaceutical production), agriculture (e.g., grain analysis), or environmental pollutant detection (2–4). More recent applications of mid-IR imaging and spectroscopy aim to establish label-free tissue assays in biological samples for medical/surgical applications, including histopathology; however, strong optical absorption by water in the mid-IR severely restricts light penetration, making biological tissues and other aqueous media effectively opaque (5–7). Sample opacity in the mid-IR spectral range restricts conventional transmission spectroscopy to excised and sliced samples, while reflection-mode (ATR) FTIR spectroscopy limits sample analysis to superficial layers (3 - 5 µm in depth), preventing access to skin compartments where dermal biomarkers, such as blood glucose, reside (8,9). Overcoming opacity and superficial sample probing is crucial to enable the development of compact mid-IR devices that can operate as biosensors across multiple anatomical sites (10).

To facilitate histopathological essays of unprocessed tissues and intravital biosensing based on vibrational spectroscopy and imaging, mid-IR optoacoustic (OA) microscopy has been developed, where light absorbed by the sample can be indirectly measured from ultrasound signals generated upon light irradiation (11,12). Initial studies involving mid-IR OA spectroscopy and microscopy systems have demonstrated living-cell imaging capabilities for cancer therapy monitoring, and biosensing potential, namely non-invasive blood glucose sensing (13–22). Especially, depth-selective sensing grants non-invasive access to information on the blood sugar in the capillary blood vessels of the human skin (10,23). However, towards establishing non-invasive biosensors based on mid-IR optoacoustics, a key limiting factor is the transmission-mode sensor configuration, where the generated ultrasound is detected from the trans-corporeal side of the mid-IR excitation due to aqueous ultrasound coupling media, otherwise blocking the mid-IR excitation light (24). Such optoacoustic sensors are thus limited to a few measurement sites on the human body, namely those that provide sufficient transmission of the light-induced ultrasound waves, for instance, the earlobes. Transmission mode sensor geometries, combined with bulky transducers for ultrasound detection, hinder the establishment as miniaturized portable biosensors.

As an alternative to optoacoustics, optothermal (OT) sensing technologies facilitate the detection of biomolecular signatures through laser-induced heat and subsequent probe beam deflectometry (25). Mid-IR optothermal sensing surpasses the need for transmission mode sensor geometries, as the signal detection can be accommodated at the illumination site because heat signal detection does not require aqueous coupling media. Recently published mid-IR optothermal spectroscopy systems have achieved measuring blood sugar in humans with errors below 10 mg/dL against reference glucose measurements (26–29). However, OT sensing demands a precise spatial arrangement of both the excitation and probe beam relative to the detector diode, where variable laser beam pointing causes optical misalignment and leads to unstable sensor readouts. Also, a major obstacle for in-vivo mid-IR biosensing is changing contact pressure and individual skin variability between humans, causing a lack of comparability of spectral measurements across different measurement sessions (30). Such low measurement reproducibility across different sessions generally leads to inconsistent signal intensities that require intensive calibration accompanied by invasive reference measurements, rendering the application of previously proposed methods as a non-invasive biosensor impractical (10,31–33). Thus, although providing a compact detector design suited for deep tissue sensing independently of anatomical measurement sites, current OT sensors lack robustness against optical instability and inter-patient skin heterogeneity, leading to poor repeatability, rendering metabolite measurements unreliable (34,35).

In order to address the lack of tissue comparability and to enhance the reliability of optothermal measurements, we developed a Spatial Probing and Autocorrection (SPAC) method for pump-and-probe mid-IR optothermal spectroscopy (OTHES) termed SPAC-OTHES. SPAC-OTHES combines 1) camera-based signal detection for spatial probe beam detection to provide readout stability against beam pointing instability and 2) self-correction using automated referencing based on superficial skin layers to compensate for varying environmental influences and skin heterogeneity on readouts from deep skin layers.

We hypothesized that SPAC-OTHES could increase analytical accuracy compared to conventional diode-based detection and provide skin-type-agnostic comparability of intravital readouts. We first tested SPAC-OTHES on phantom measurements and subsequently conducted in vivo measurements on healthy volunteers to assess SPAC-OTHES reproducibility and the comparability of spectral measurements of human skin. Our results demonstrate that measuring the spatial probe beam deformation yields lower glucose quantification errors in skin-like phantoms compared to conventional diode-based deflectometry. Moreover, we demonstrate that self-reference correction facilitates spectral stabilization and improves comparability of measurements across different human volunteers and on different days.

## Results

### Working principle of SPAC-OTHES

SPAC-OTHES is a pump-and-probe mid-IR spectroscopy system based on all-optical detection of optothermal signals generated by mid-IR excitation. **Fig. 1a** shows a schematic representation of the system, with details in the **Methods** section. Briefly, in SPAC-OTHES, a customized optical component, termed optothermal window (OT-window), in direct contact with the sample of interest, is configured to efficiently allow a modulated mid-IR beam (pump beam) to irradiate the sample while simultaneously reflecting a coaligned interrogating beam (probe beam) at the surface directly adjacent to the sample-window interface on the mid-IR irradiating area. As the sample absorbs and, subsequently, releases the absorbed mid-IR energy in the form of heat (the optothermal effect), a localized temperature gradient diffuses from the sample to the OT-window, changing its refractive index and thus perturbing the reflected probe beam. The system was designed to operate in a co-aligned pump- and probe-beam optical configuration to facilitate future hardware miniaturization by reducing the complexity of optothermal detection. The perturbation of the probe beam is simultaneously retrieved over time in two ways: 1) by a photodetector, as typically done in OT sensing, and 2) uniquely to SPAC-OTHES by a camera that monitors the spatial distribution of the perturbed probe beam profile, with which we expect to achieve an improved readout stability compared to diode detection (i.e., using quadrant diodes).

**Fig. 1.**
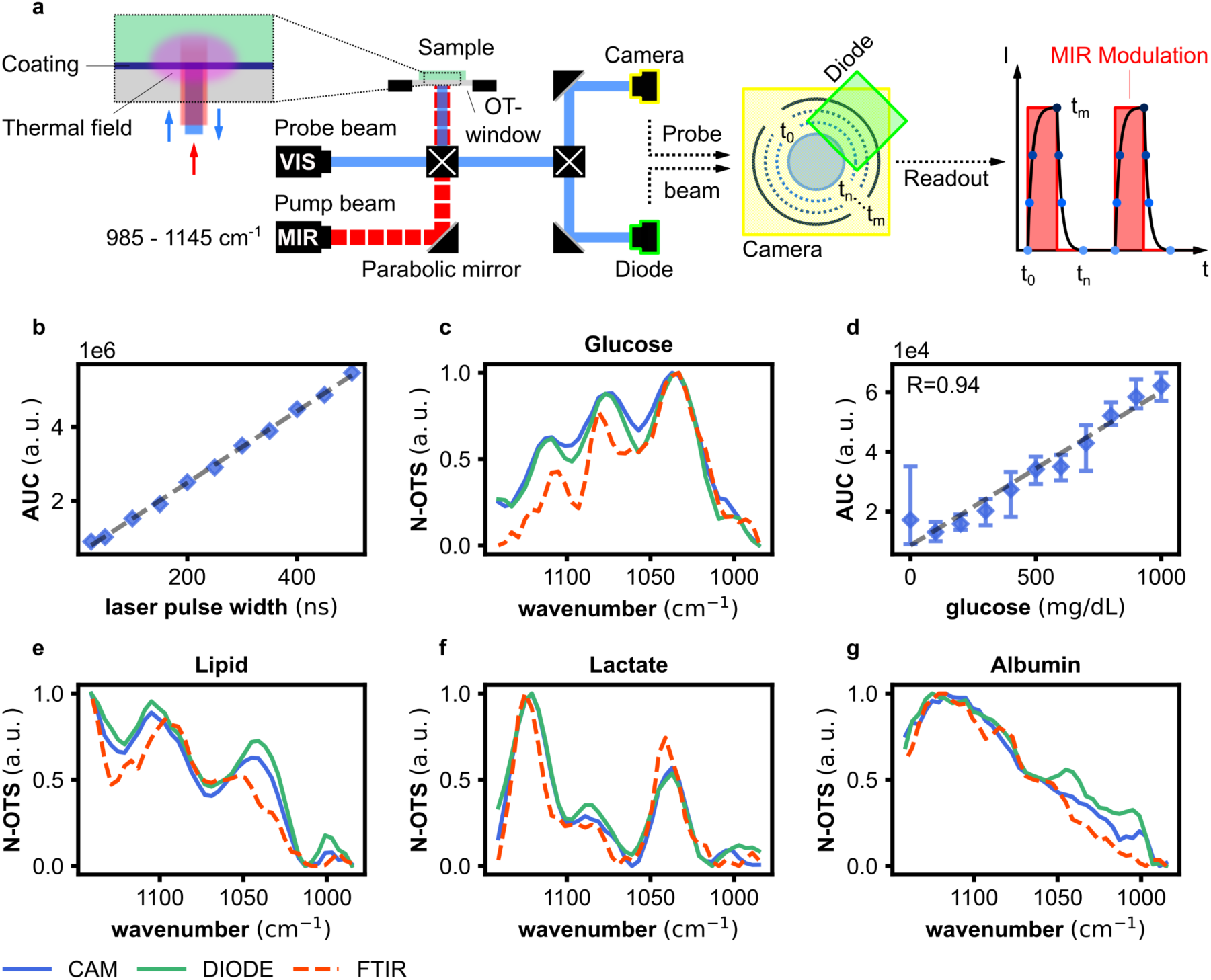
Mid-infrared optothermal pump-and-probe spectroscopy. **a,** Working principle of Spatial Probing and Autocorrection Optothermal Spectroscopy (SPAC-OTHES). The system consists of a mid-infrared (MIR) quantum cascade laser (QCL), which irradiates a sample to induce the optothermal effect. The optothermal effect generates a heat field in the sample as well as the sample window, which is probed by a visible (VIS) laser. A camera and a diode assess the reflected beam from the VIS laser to quantify the intensity of the optothermal response and, thus, the MIR absorption in the sample. Depending on the signal acquisition delay, different depth slices can be probed. **b**, Signal detection linearity. The graph shows the areas under the curve of SPAC-OTHES intensity profiles of a water sample with varying laser power levels. **c**, Comparison of glucose absorption spectra retrieved by SPAC-OTHES with Fourier transform infrared spectroscopy (FTIR) spectra. **d**, SPAC-OTHES spectra of water-glucose phantoms with glucose concentrations varying from 0 to 1000 mg/dL. The graph shows the areas under the curve of SPAC-OTHES spectra. **e**, Comparison of lipid absorption spectra retrieved by SPAC-OTHES with FTIR spectra. **f**, Comparison of lactate absorption spectra retrieved by SPAC-OTHES with FTIR spectra. **g**, Comparison of albumin absorption spectra retrieved by SPAC-OTHES with FTIR spectra.

As schematically represented in **Fig. 1a** and shown in **Supplementary Fig. 1a** for water, as expected, signal readouts based on probe beam detection using a photodiode revealed a periodic probe-beam intensity response in sync with the cycles of the modulated mid-IR excitation. The temporal behavior of the detected signal corresponds to the typical heat-up/cool-down cycles of OT detection. When employing camera-based spatial probe beam detection, we observed that the intensity changes of the probe-beam profile also retrieved the heat-up/cool-down cycles, as shown in **Supplementary Fig. 1b,** like the response of the photodetector. The spatial probe-beam detection revealed shape changes predominantly occurring as beam defocusing. Importantly, both diode and camera detection show a linear OT signal response with increasing mid-IR laser power, see **Fig. 1b**.

In SPAC-OTHES, a mid-IR optothermal spectrum (OTS) is obtained by recording the OT intensities at individual wavenumbers in the range of 985-1145 cm^-1^ and subsequently correcting for the wavelength-dependent pump laser emission power. The spectroscopic abilities of SPAC-OTHES, retrieved by diode and camera detection, are demonstrated in **Fig. 1c**, where the OTS of glucose is compared to the mid-IR spectra of the same sample obtained by standard Fourier-transform infrared spectroscopy (FTIR). We observed that SPAC-OTHES and FTIR spectroscopy reproduce overly matching spectral features of glucose, **Fig. 1c**. Additionally, OTS of glucose-water solutions measured at different concentrations, in the range between 0 and 1000 mg/dL, reveals a linear response of the OTS with molecular concentration; a crucial characteristic for glucose sensing, see **Fig. 1d**. Similar results (i.e., spectral accuracy and linearity with concentration) were confirmed in other important biomolecules commonly found in biological tissues, see for instance **Figs. 1e-g** for intralipid, lactate, and albumin.

### Quantitative glucose sensing in liquid skin phantoms

Next, beyond simple single-analyte solutions, we tested the accuracy of SPAC-OTHES in complex heterogeneous analyte solutions mimicking a mixture of biomolecules typically found in the shallow layers of skin—termed liquid skin—and compared measurement performance when using a camera and a photodiode for OT detection. For this purpose, we generated a total of 80 liquid skin phantoms at varying concentrations of Intralipid, lactate, and albumin in water, simulating the molecular variability of different skin types from patient to patient; each at ten different glycemic levels ranging from 0 to 800 mg/dL (see **Methods**). In each measurement, the modulated OT signal was recorded simultaneously with the camera and the diode to ensure direct comparability. To quantify the glucose concentrations in the liquid-skin phantoms, baseline calibration followed by glucose prediction using partial least squares regression (PLSR) was applied in an 8-fold cross-validation scheme. In baseline calibration, a spectrum of the liquid-skin specific molecular composition is subtracted from each spectrum of the same skin type to compensate for the molecular baseline, as described in **Methods. Fig. 2a** visualizes the glucose quantification scheme, where for each liquid skin phantom, glucose concentration was predicted after training PLSR models based on the spectra of the remaining liquid skin phantoms. The modeling approach with skin-type-based cross-validation ensures that the training and test sets contain independent samples by minimizing confounding factors in both datasets. In this way, the retrieved error metrics are expected to reflect the quantitative glucose detection capability of SPAC-OTHES for each newly measured phantom, independent of skin composition.

**Fig. 2.**
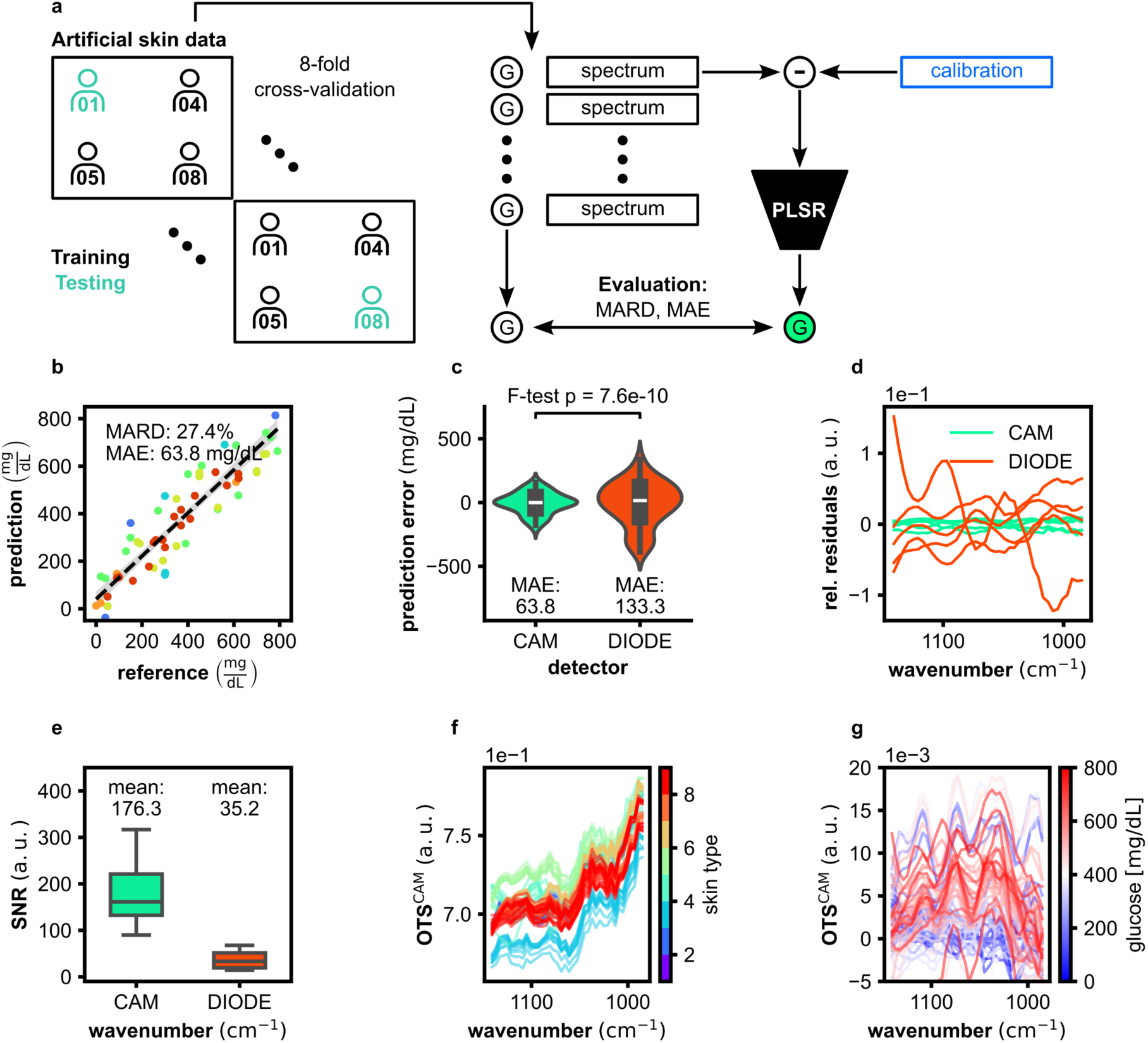
Glucose quantification in artificial skin phantoms. **a,** Artificial skin phantom measurements using SPAC-OTHES. A total of 8 artificial skin types were created and superimposed with 10 different glucose concentrations in each skin type. In this way, a dataset containing 80 molecular compositions could be used for training quantification models using leave-one-skin-type-out cross-validation. **b**, Glucose prediction results based on partial least squares regression (PLSR). **c**, Statistical error analysis comparing camera-based and diode-based glucose quantification using PLSR. **d,** Relative residuals of longitudinally repeated measurements comparing diode- and camera-based signals readout. **e**, Signal-to-noise ratios (SNRs) of SPAC-OTHES spectra retrieved using camera-based versus diode-based signal detection. **f**, SPAC-OTHES spectra of all artificial skin phantoms retrieved via camera detection, which are color-coded by the artificial skin type. **g**, Calibrated SPAC-OTHES spectra of all artificial skin phantoms retrieved via camera detection, which are color-coded by glucose concentration.

The comparison of quantification errors obtained via camera and diode-based glucose quantification reveals that camera-based detection delivers significantly higher accuracy than diode-based detection (F-test p-value < 0.05). Namely, camera-based detection leads to a MAE of ca. 64 mg/dL while diode-based detection has a MAE of 133 mg/dL, as shown in **Figs. 2a-b**. The remarkable differences in the ability of both detection methods for glucose quantification on liquid skin phantoms can be explained by the large differences in measurement stability and SNR between the two readout methods observed, supporting the hypothesis of improved robustness by camera-based detection in comparison to photodiode detection.

The differences in measurement stability were assessed by testing the robustness of camera and diode-based detection in liquid skin phantoms. For this purpose, identical phantom solutions were measured and compared regarding their repeatability. The relative repetition residuals, i.e., the differences between the repeatedly measured spectra relative to the spectral intensity, show for the camera distinctively lower values indicating better repeatability than the residuals corresponding to the diode-based detection, as shown in **Fig. 2c**. A comparison of the camera and diode detection results on the liquid skin phantoms revealed that the camera-based detection leads to a mean SNR of ca. 176, which is approximately 5 times higher than for diode detection, which has a SNR of ca. 35 as displayed in **Fig. 2d**. **Figs. 2e-g** show the raw camera-based OTS, color-coded by the different baseline compositions, and the OTS after baseline calibration, color-coded by the glucose concentration, respectively. The comparison between the two visualizations shows that the measured spectra primarily reflect the difference between artificial skin types rather than glucose levels, highlighting the necessity for a method to increase spectral comparability between different skin types.

Our results show the advantage of camera-based detection for measurement repeatability over diode-based detection, which is especially relevant moving towards in vivo application. In combination with a significantly higher SNR, camera-based detection was able to achieve improved glucose quantification results in phantoms with complex molecular compositions compared to classical photodiode-based detection.

### Depth measurements on multi-layer samples

An important characteristic of OT spectroscopy is the ability to obtain spectral information at different depths—termed spectral depth profiling—in a sample. Spectral depth profiling is achieved by varying signal acquisition delays (i.e., time gating) as described in **Methods**, since the deeper the OT signal is generated inside the sample, the longer it takes for the generated heat to arrive at the sample surface and OT window. Here, we demonstrated the optothermal spectral depth profiling ability of SPAC-OTHES by acquiring spectra at different depths on a customized PTFE-paper phantom. PTFE and paper have remarkably different spectral features that make it easy to distinguish between each other, see **Fig. 3a**. The investigated phantom was set to have a well-defined PTFE layer thickness (50 μm in this example) in direct contact with the OT-window and a sheet of paper on top of the PTFE layer, see **Fig. 3b**. As the OT signal is acquired at twelve different time arrivals, from early to late arrival, we observed how the obtained phantom spectra transitioned from that of PTFE on the surface to the one of paper located deeper in the phantom, see **Fig. 3c**. In the representative example in **Fig. 3b**, SPAC-OTHESHES demonstrated the ability to retrieve mid-IR spectra beyond the 50 µm PTFE layer. Moreover, reaching deeper layers, up to 100 µm, was demonstrated by increasing the PTFE thickness, see **Supplementary Fig. 2**, although at a lower SNR compared to the measurement with the 50 µm layer at an acquisition delay of 24 ms.

**Fig. 3.**
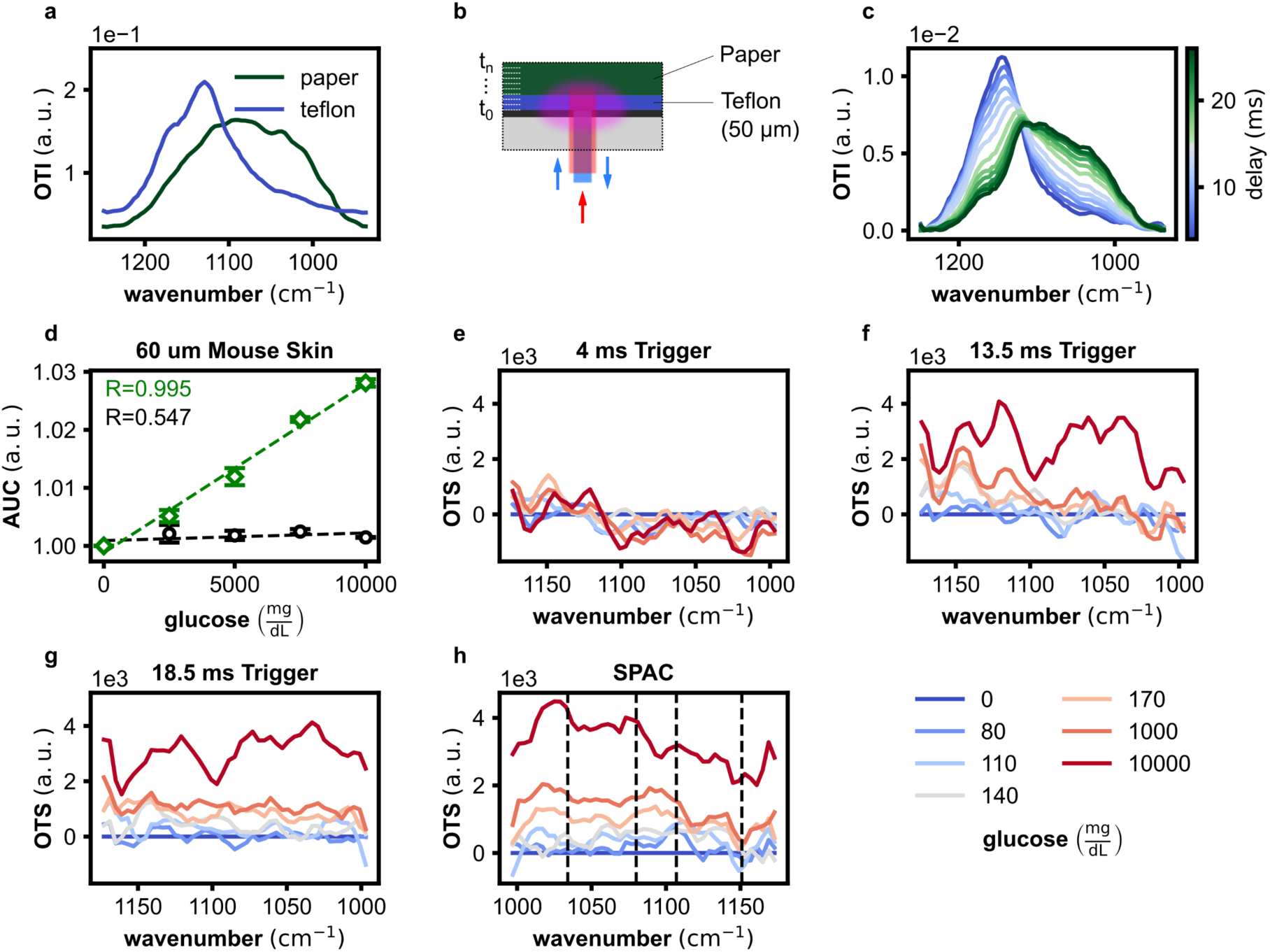
Depth selective optothermal measurements of multi-layer samples. **a**, Intensity profiles of a PTFE and a paper sample measured with our optothermal system. **b**, Schematic representation of a layered phantom. The phantom consists of a 50 μm PTFE layer with a paper sample placed on top. **c**, Depth signal of the layered phantom. Different acquisition delays enable the retrieval of signals from different depths of the layered phantom. **d**, Relative change of the area under the curve for measurements of a multi-layer sample consisting of 60 µm mouse skin, 10 µm PE foil, and 150 µL water with glucose concentrations from 0 – 10000 mg/dL. The measurements were performed with a signal acquisition delay of 13.5 ms and 19.5 ms. **e-g**, Change of system response of a similar multi-layer sample with a 40 µm mouse skin layer and glucose concentrations from 0 – 10000 mg/dL separated by signal acquisition delay. **h**, Self-calibrated change of system response in which the measurement with a 4 ms acquisition delay was used to correct the data for spectral changes of the mouse skin layer.

After showing the capabilities of SPAC-OTHES for spectral depth profiling, we used a 60 µm-thick cryosectioned mouse skin to investigate the advantages of depth measurements for metabolite sensing beyond an initial layer of skin. For this purpose, we placed water solutions with different glucose concentrations on top of the skin sample, separated from the skin by a 10 µm thin layer of PE foil to prevent diffusion, and measured them with SPAC-OTHES. **Fig. 3d** displays the relative magnitude change of the system response with increasing glucose concentration measured at the end of the laser irradiation period with an acquisition delay of 13.5 ms (black) and 6 ms later, after 19.5 ms during the cool-down phase of the sample (green). The results demonstrate the ability of SPAC-OTHES for glucose sensing beyond an initial skin layer with high linearity (R=0.995) by delaying the data acquisition to a timepoint in the cool-down phase of the sample and therefore increasing the measurement depth. In contrast, data acquisition at the point of maximum irradiation led to a higher signal and SNR (566 at 13.5 ms compared to 261 at 19.5 ms) but was not able to detect intensity changes correlated to the glucose concentration of the sample. With this experiment, we were able to prove that spectral depth profiling enables glucose quantification beyond a skin layer of 60 µm. However, we also discovered that the skin layer, like human skin, introduces a measurement instability due to absorption changes over the duration of the experiment, which negatively impacts the ability to isolate glucose-specific changes.

The ability of SPAC-OTHES to capture sample-layer-dependent information, depending on the acquisition delay, enabled the implementation of a new method for self-corrected measurements to address the measurement instabilities associated with biological samples. Self-correction is made possible by simultaneous data acquisition from multiple depths, including at least one superficial measurement, by performing data acquisition at delays of 4, 13.5, and 18.5 ms in relation to the start of each mid-IR irradiation cycle. The absorption data from the shallow layer is then used to correct the glucose-relevant measurement of deeper depths. Self-correction was evaluated in measurements of 40 µm mouse skin and water droplets of increasing glucose concentration, similar to the setup described in the previous section. Displayed in **Figs. 3e-g** are the system responses for the different glucose concentrations separated by acquisition delay, hence depth. The data acquired with the acquisition delay of 4 ms, resulting in a shallow measurement, shows no correlation to the glucose concentration, as expected based on the previous findings. In contrast, the system responses captured with an acquisition delay of 13.5 and 18.5 ms both display clear changes in magnitude depending on the glucose concentration. However, even for very high glucose concentrations, i.e., 10,000 mg/dL, it was not possible to extract the characteristic glucose absorption peaks due to the superimposed changes of the skin sample over the duration of the experiment. The superficial data acquired with a delay of 4 ms captures exactly these changes of the skin sample, isolated from the glucose information, enabling self-correction by filtering out non-glucose-related changes from the measurements of deeper depths. The SPAC-OTHES data displayed in **Fig. 3h** demonstrates the possibility of retrieving the glucose-specific absorption peaks using self-correction from the same measurement for high glucose concentrations indicated by the dashed lines. Correcting measurements from deeper sample layers with the superficial measurement enables the exclusion of measurement instabilities caused by the instability of biological samples, which becomes especially relevant in the translation to in vivo measurements.

### SPAC-OTHES for measurement stability and inter-person comparability

Moving towards self-corrected in vivo measurements, we applied optothermal spectroscopy with time gating in 20 healthy human volunteers to investigate the spectral comparability between different skin types, measurement days, and optical alignments, comparing the default single depth measurement (13.5 ms acquisition delay) with the SPAC-OTHES self-correction method (collection of all measurements in **Supplementary Fig. 3 and 4**). For each volunteer, 25 measurements were performed over a period of two hours while monitoring their glycemic level with conventional invasive blood-glucose measurements (see **Methods**). As expected, volunteer movement and the resulting pressure changes on the sample window introduced minimal changes in the reflection angle, resulting in a pointing instability of the probe beam over the 2h measurement period, as displayed in **Supplementary Fig. 5**. The maximum probe beam displacement over the period of one OGTT was, on average, 95 µm in the volunteer study. In comparison, the probe-beam drift in the measurements of the multi-layer mouse skin measurements was, with an average of 11 µm, more than eight times lower, highlighting the impact of volunteer movement introduced by pointing instabilities. An analysis of the beam profile showed that a shift of 95 µm away from the center of the probe beam can lead to a signal loss up to 25% and thus introduce high measurement instabilities in diode-based detection. With SPAC-OTHES, the whole beam area is captured at all times, which decreases signal instabilities related to probe-beam drift caused by volunteer movement, resulting in an average SNR of 330 for camera-based detection and 73 for diode-based detection.

**Fig. 4.**
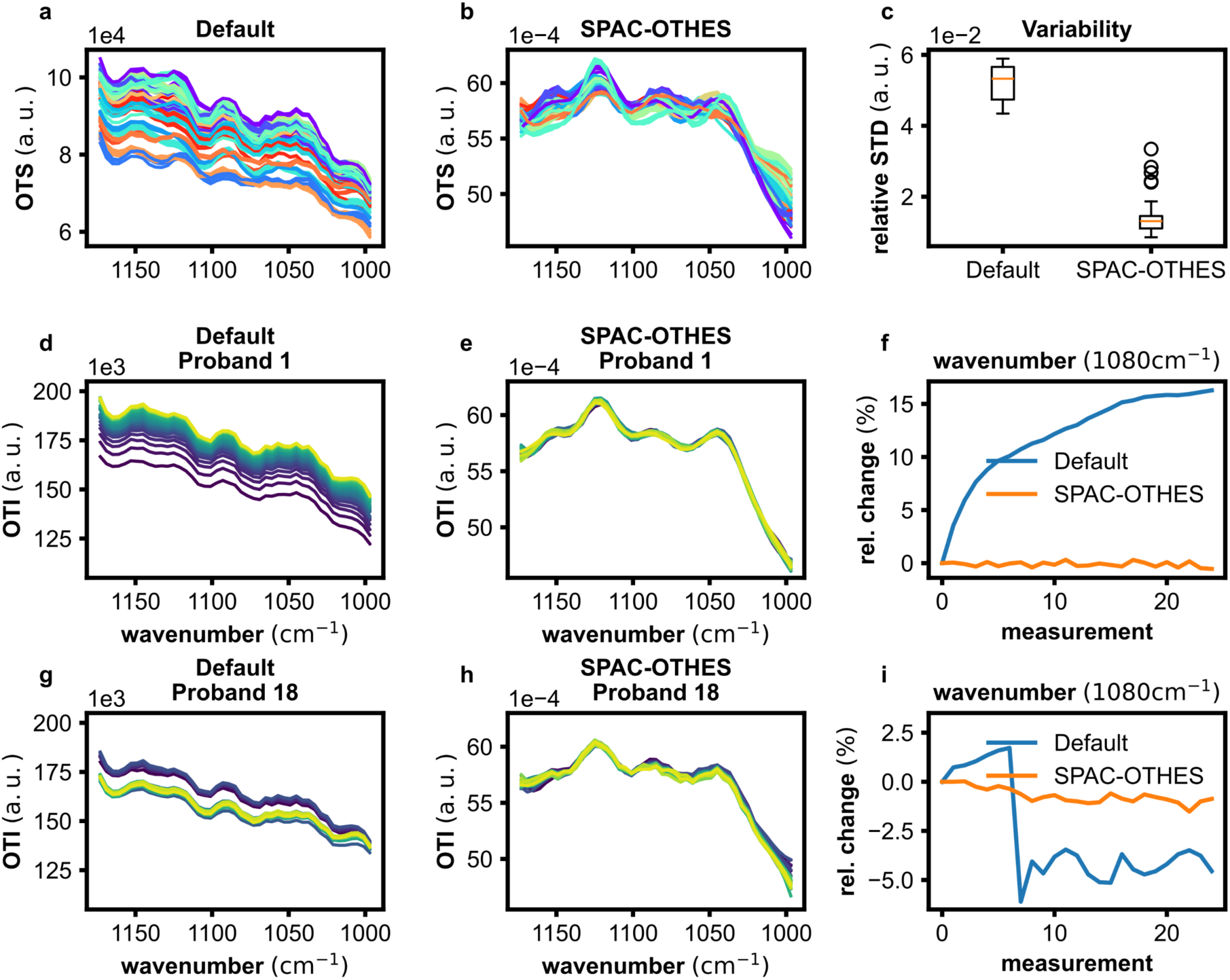
SPAC-OTHES for robust intra-vital mid-infrared spectroscopy. **a**, 100 Spectra of 20 human volunteers with an average blood glucose level of 154.2 ± 6.8 mg/dL captured at the end of the mid-IR irradiation period, termed default, measured on different days, and colored by individual volunteer. **b**, Data from the same measurements as in **a**, processed with autocorrection. **c**, Comparison of the relative standard deviation over all wavelengths between the default and autocorrected data. **d**, 25 default measurements from volunteer 1 recorded over a time of two hours, colored by measurement time. **e**, Autocorrected measurement data from volunteer 1, colored by measurement time. **f**, Comparison of the relative signal magnitude change over the 25 measurements between the default and autocorrected data on the example of a single wavenumber. **g**, 25 default measurements from volunteer 18 recorded over a time of two hours, colored by measurement time. **h**, Autocorrected data from volunteer 18 colored by measurement time. **i**, Comparison of the relative signal magnitude change over the 25 measurements between the default and autocorrected data on the example of a single wavenumber.

**Fig. 4a** shows 100 scans acquired from 20 different volunteers on 20 different days with comparable metabolite concentration (± 6.8 mg/dL glucose), colored by volunteer number, displaying a high variance of system response not related to metabolite concentration. Clear spectral separation is observable, especially between different volunteers, caused by differences in, for example, skin type, sensor contact, and optical setup. We designed SPAC-OTHES to facilitate inter-person comparability of spectral readouts independent of volunteer and measurement day. As a foundation for auto-correction, we used spectral depth profiling as introduced in the previous section and three different acquisition delays at 4, 13.5, and 18.5 ms after the start of the mid-IR irradiation period. The shallow measurement after 4 ms was used to filter out changes of the superficial skin over time by correcting the changes in the measurements of deeper depths by the changes of the 4 ms measurement and thus stabilizing the recorded system response. To reduce the influence of skin-specific absorption, which differs significantly between different volunteers, we used the self-corrected 13.5 and 18.5 ms system responses to calculate the slope of the cool-down curve instead of analyzing them individually, thus isolating spectral data coming from deeper depths beyond the initial stratum corneum. **Figs. 4b,c** show the potential of SPAC-OTHES to increase spectral comparability by significantly reducing the impact of confounding factors, i.e., skin contact pressure, skin-type, and optical alignment in in vivo measurements. SPAC-OTHES enables a decrease of the relative standard deviation over all wavenumbers from 5.2% to 1.4%, which is a reduction of 73%, demonstrating a significant increase in inter-person comparability and measurement stability.

In **Figs. 4d-f**, we show the unique ability of SPAC-OTHES to compensate for the measurement of arbitrary signal drift over time, on the example of volunteer 1, enabled by the correction of the data with the isolated signal drift in the superficial measurement. **Fig. 4f** displays the signal drift on the example of one wavenumber, which exhibits a relative change of 16.3% over the measurement duration of two hours, while SPAC-OTHES reduces the relative change to 0.5% in the same period. Another advantage of SPAC-OTHES is the correction of measurement location and contact pressure-related signal changes. For instance, volunteer 18 was asked to remove their wrist from the sensor and then place it back on the sensor to continue the measurement. **Figs. 4g-i** display the change in signal magnitude 30 min after the start of the volunteer measurement, corresponding to the readjustment of wrist position and contact pressure captured with the single depth measurement labeled as default. The probe-beam center shifted position by 142 µm during the duration of the measurement. SPAC-OTHES enables spectral correction, which is designed to overcome measurement instabilities demonstrated in **Figs. 4h-i**. A relative signal loss of 7.8% was recorded between the two measurements before and after the wrist reposition using single depth measurement, while SPAC-OTHES self-correction reduces the signal loss to 0.3% in the same period. Altogether, the results of the volunteer study show the potential of SPAC-OTHES to enable inter-person comparability between different days and optical alignments.

## Discussion

We demonstrated that SPAC-OTHES enables robust intravital mid-IR spectroscopy at depths (>60 µm), far beyond the limits of conventional mid-IR ATR spectroscopy (<5 µm). In particular, in phantoms, spatially-resolved probe-beam OTHES detection achieved up to 52% lower glucose quantification errors compared to photodiode-based detection. Furthermore, we showed that auto-calibrated spectral measurements by skin-layer-based self-referencing correct for patient-to-patient and day-to-day variability across different probands and measurement conditions, which results in 73% lower spectral fluctuations in intravital measurements of human probands compared to non-corrected readouts.

SPAC-OTHES is the first optothermal spectroscopy system to capture spatial data to enable measurement stabilization. Experiments on liquid skin phantoms revealed that SPAC-OTHES can quantify glucose in complex and heterogeneous skin-like phantoms. Using camera-based detection combined with computational signal processing using PLSR, we observed an overall detection accuracy given by a MAE of approximately 64 mg/dL; ca. 52% lower compared to photodiode-based detection. In photodiode-based detection, the diode surface is aligned to a small proportion of the probe beam, which fluctuates upon optothermal effect induction, while the camera instead captures the whole beam and is thus able to compensate for beam pointing instabilities. This compensation for pointing instability is also reflected by a lower SNR for single-point detection, i.e., ca. 35, instead of 176 arbitrary units for camera-based detection. Since most optothermal pump-and-probe spectroscopy sensors use photodiode (quadrant-diode) detection, this finding can notably improve the fidelity of any mid-IR optothermal sensing system when adjusting the detection mechanism accordingly. Although the absolute error for glucose sensing must be improved approximately by an order of magnitude to ensure regulatory approval as an intravital glucose sensing device, we anticipate substantial improvements from further technical improvements to reduce beam pointing instabilities, such as, for instance, the use of active beam steering for the QCL (36).

The measurements of the layered PTFE-paper phantom show that SPAC-OTHES can probe layers beyond 50 μm in opaque samples. Conventional mid-IR spectroscopic methods for measuring opaque samples often use Attenuated-Total internal Reflectance (ATR) configurations, for instance, ATR-FTIR. These technologies can only probe superficial layers (2-5 μm in depth), which limits their application as biosensors for humans since important biomarkers, including glucose, reside in skin layers beyond 20 μm (37). Additionally, contrary to depth-resolved mid-IR OA spectroscopy, where detection is achieved by bulky transmission-mode excitation/detection, SPAC-OTHES detection is achieved in compact reflection-mode, which facilitates application independently of anatomic location (38,39). Similar to prior OT sensing devices, using frequency-modulated layer-selection, SPAC-OTHES can measure in reflection mode and, at the same time, retrieve signals from varying sample layers; however, beyond frequency-modulated layer-selection, layer-selection in SPAC-OTHES is tied to the illumination time after the laser pulse onset, enabling simultaneous measurement of different depths (40). For the first time in OT spectroscopy, we used the principle of retrieving signals from different sample layers to isolate superficial spectra to identify measurement instabilities and calibrate signals retrieved from the deeper layers. In SPAC-OTHES, this calibration is achieved by a reference signal acquired using a short delay of 4 ms after the start of the mid-IR irradiation to stabilize measurements from layers with relevance for intravital glucose sensing. The ability of measurement stabilization was subsequently transferred to the in vivo measurements.

SPAC-OTHES acquires and processes OT signals from multiple depths simultaneously and therein facilitates self-correction to enable inter-personal comparability of spectral measurements. To make measurements comparable with respect to different measurement sessions and probands, SPAC-OTHES uses the outermost skin layer for real-time signal calibration. The results based on the in-vivo experiments in human probands show, in comparison to non-calibrated measurements, that SPAC-OTHES achieves a reduction of the relative standard deviation of spectral readouts from 5.2% to 1.4%, which corresponds to a reduction of spectral variations by 73%. It follows that SPAC-OTHES is able to compensate for varying measurement and proband skin characteristics across different measurement sessions. Besides often insufficient spectral scaling and normalization, most methods in the literature fail to provide specialized mechanisms to compensate for effects associated with varying skin properties across probands, varying contact pressure, and alignment drift. However, missing measurement stabilization mechanisms can lead to low prediction fidelity or failure to establish data processing methods for glucose quantification that generalize for a large proband cohort. For instance, many studies trained personalized models, which led to reasonable quantification errors for single probands but failed to establish quantification for temporally independent measurement sessions and probands (38,39,41–46). However, the fidelity of glucose quantification by personalized processing models cannot be sustained in real-world applications due to a lack of generalizability without impractical calibration outside laboratory conditions. In contrast, SPAC-OTHES uses built-in measurement self-correction for real-time spectral stabilization, setting the stage for generalized calibration-free metabolite quantification in future developments.

Overall, SPAC-OTHES provides a compact, normalized intravital mid-IR spectroscopy system, which achieves better stability against optical alignment drift than conventional photodiode detection. In this way, it enables auto-calibration and correction for varying measurement conditions and different probands.

## Methods

### Pump-and-probe optical setup

The developed optothermal spectroscopy system uses a pump-and-probe detection principle, where an external cavity quantum cascade laser (EC-QCL Hedgehog, Daylight Solutions) with a tuning range from 8-10.6 µm is focused with a parabolic mirror (MPD169-M03, Thorlabs) and directed onto the sample for mid-IR excitation. The laser was operated with a current of 950 mA and a pulse width of 500 ns, except for the linearity study with changing pulse width as specified in the **System characterization experiments** section. A waveform generator (DG1022, Rigol) provides a 100 kHz signal to trigger the laser pulses. The QCL is digitally modulated using a microcontroller (ESP32, Espressif) to generate modulation periods during which the laser emits pulses over 13.5 ms, followed by 270.3 ms of no emission, creating heat-up and cool-down cycles that enable thermal diffusion from the sample into the window. The QCL is tuned from 937 cm^-1^ to 1249 cm^-1^ in steps of 4 cm^-1^ while retaining each sampled wavenumber over three modulation periods before tuning it to the next wavenumber to enable data averaging for noise reduction. However, only the wavelengths between 985 cm^-1^ and 1145 cm^-1^ were used to compute the final spectra since beyond these bounds the QCL has too low emission intensity to enable generating stable optothermal signals. The probe laser, a single-wavelength continuous wave laser (iBeam smart 405 S-LP, Toptica) with a wavelength of 405 nm, is operated at 90 mW, sent through a ND1 filter, and pointed to the spot illuminated by the QCL using a beam combiner. The sample is positioned on a dichroic mirror coated to transmit the mid-IR beam and to reflect the visible beam on the surface facing the sample. The reflected probe laser is directed through two beam splitters (BS013, Thorlabs) to capture it with a camera (Flea3 FL3-U3-20E4M, Point Grey Research) and a diode (DET10A/M, Thorlabs). The camera captures images on an external trigger provided by the microcontroller at the start and the end of each modulation period of the QCL. A DAQ (cDAQ-9174, National Instruments) collects the transient diode and QCL modulation signals to enable subsequent processing of the diode data based on the on-off timing of the laser. The QCL, the microcontroller, the camera, the diode, and the DAQ are controlled via a Python script, enabling automation and precise data capturing and processing.

### Signal processing for retrieval of optothermal spectra

To generate optothermal spectra, the camera-based data and the diode-based data are processed separately via a Python script. In the camera-based approach, image pairs representing the on and off states of the QCL are collected three times for each sampled wavenumber. Thereby, each image pair corresponds to one of the three modulation periods. In the next step, the corresponding image pairs were subtracted from each other, resulting in three images 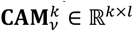 to be averaged for each wavenumber 𝜈, where 𝑗 ∈ {1; 2; 3} and 𝑘, 𝑙 are the pixel dimensions. Based on the difference images, the intensity values OTI*_v_* are computed as

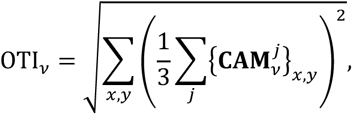

where 𝑥, 𝑦 are indexing the camera pixels. Similarly, the diode data is processed for each wavenumber individually, where the transient signal retrieved by the DAQ is separated into three intervals corresponding to the modulation period of the QCL and resulting in three heat-up and cool-down curves 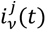, where 𝑗 ∈ {1; 2; 3}. For diode-based sensing, the corresponding intensity values are computed as

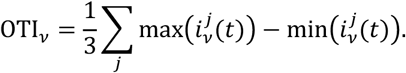

In total, 𝑛 intensities OTI_!_ representing the wavenumbers 𝜈_1_ … 𝜈_𝑛_ (985 cm^-1^ and 1145 cm^-1^) are then concatenated and subsequently filtered using a Savitzky-Golay filter, resulting in the optothermal intensity profiles 𝐎𝐓𝐈 ∈ ℝ^𝑛^. Filtering is employed to reduce the displacement effects of the mid-IR laser beam relative to the probe beam, since most quantum cascade lasers (QCLs) have pointing and beam shape instabilities over different wavelengths (47,48). Furthermore, the pointing instability can be reduced via a parabolic mirror in the optical setup, which slightly focuses the mid-IR beam. For default processing without autocorrection, the optothermal spectra are obtained by element-wise division of 𝐎𝐓𝐈 by the emission intensity profile of the QCL.

### Autocorrection

For autocorrection of the spectra acquired using camera-based detection, three intensity profiles 𝐎𝐓𝐈_𝑡_ are captured quasi-simultaneously in each measurement, each corresponding to an acquisition delay of 𝑡 = 4 ms, 𝑡 = 13.5 ms, and 𝑡 = 18.5 ms relative to the start of the mid-IR irradiation. The autocorrection mechanism consists of two consecutive steps:

1. **Drift compensation:** To correct signal drift across different measurement sessions (and thus different study probands), all measured intensity profiles 𝐎𝐓𝐈_𝑡_ are referenced with the change of 𝐎𝐓𝐈_4 ms_ over time. Therefore, the signal change of the 𝐎𝐓𝐈_𝑡_ over one measurement session is first determined as

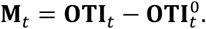

Since a change in the relative acquisition delay results in a change in signal magnitude, as displayed in **Supplementary Fig. 1**, 𝐌_𝑡0_ and 𝐌_𝑡1_ (𝑡_0_, 𝑡_1_ ∈ {4 ms, 13.5 ms, 18.5 ms}, 𝑡_0_ ≠ 𝑡_1_), cannot be directly compared without intensity scaling. To achieve scaling of intensities associated with different trigger delays and at the same time compensate for drift, a calibration operator, 𝐹: ℝ^𝑛^ → ℝ^𝑛^, is determined. Thereby, 𝐹 is assumed constant across one intensity profile and chosen as a linear operator

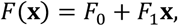

The parameter values 𝐹_0_ and 𝐹_1_ are obtained by least squared fitting of the difference between the operator applied to the measurements 𝐌_4 ms_ and later measurements 𝐌_𝑡_, i.e., 𝐌_13.5 ms_ and ^𝐌^_18.5 ms_, as 𝐹_0_, 𝐹_1_ = arg min‖𝐌_𝑡_ − 𝐹(𝐌_4 ms_)‖^2^.

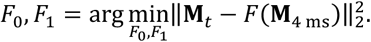

Based on the determined parameters of 𝐹, the drift-compensated intensities 𝐎^^^𝐓𝐈_𝑡_ are given as the difference between 𝐎𝐓𝐈_𝑡_ and the adjusted intensity change of the 4 ms measurement, i.e.,

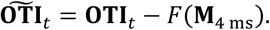

1. **Depth signal extraction:** The second step consists of extracting absorption information of deep skin layers, rejecting the absorption in the outermost layers. Therefore, the intensity profiles 𝐒𝐏𝐀𝐂𝐈 ∈ ℝ^𝑛^ are computed based on the drift compensated intensity profiles 𝐎^^^𝐓𝐈_13.5 ms_ and 𝐎^^^𝐓𝐈_18.5 ms_ and can be interpreted as a time constant given as

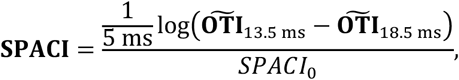

which describes the sample cool-down process after mid-IR light excitation. This cool-down process is assumed to follow an exponential decay and 𝑆𝑃𝐴𝐶𝐼_0_ denotes a normalization constant given as

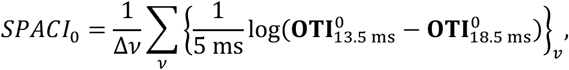

where 𝐎𝐓𝐈^0^_13.5 ms_ and 𝐎𝐓𝐈^0^_18.5 ms_ are the first measurements of a measurement session of a study proband.

Lastly, the 𝐒𝐏𝐀𝐂𝐈 are filtered by a Savitzky-Golay filter and divided by the laser emission profile of the mid-IR laser to obtain the final autocorrected spectra 𝐒𝐏𝐀𝐂𝐒 ∈ ℝ^𝑛^.

### System characterization experiments

Optothermal spectroscopy measurements were performed on high-concentration samples of four molecules present in the human skin to assess the spectroscopic capabilities of the system. Therefore, 1 mL samples of 5000 mg/dL glucose (G7528, Sigma-Aldrich), 20000 mg/dL intralipid (I141, Sigma-Aldrich), 4000 mg/dL lactate (L7022, Sigma-Aldrich), and 15000 mg/dL albumin (Art.-Nr. 8076.2, Carl Roth) were prepared with deionized water. The SPAC-OTHES system was used to measure the OTS of the molecule samples and a baseline sample of deionized water, each in five scans. Then, the molecule spectra were calculated by subtracting the water baseline from the measurement of the molecule solution. Additionally, ground truth measurements of the infrared absorption spectra of the samples were obtained with an FTIR spectrometer (Alpha II, Bruker). For the assessment of the system linearity to different glucose concentrations, eleven samples of 1 mL deionized water with increasing glucose concentration from 0 mg/dL to 1000 mg/dL were prepared in steps of 100 mg/dL. After measurement of the samples, the absorption spectrum of the 0 mg/dL sample was used to subtract the water absorption and isolate the glucose spectra in the remaining measurements. For the QCL power linearity assessment, absorption spectra of deionized water were recorded at different QCL duty cycles by changing the pulse width while retaining a constant repetition rate of 100 kHz. Thereby, measurements were obtained for each pulse width from 500 ns to 50 ns, decreasing in steps of 50 ns, including an additional measurement with a pulse width of 25 ns. The system response to the changing duty cycle was compared by calculating the area under the curve of the measurements. In a subsequent experiment, the beam wandering of the QCL was recorded with an IR camera (WinCamD-IR-BB, DataRay) above the sample window by tuning the QCL through the available wavelengths five times in total to assess reproducibility. The beam profile and intensity were captured with the DataRay software and a custom Python script, in which the center of the beam was defined as the mean of a Gaussian fit to the beam profile.

### Liquid skin phantom study

Water solutions with varying concentrations of lipid, albumin, and lactate, so-called liquid skin samples, were prepared to simulate skin-like mid-IR absorption properties. Each liquid skin type was prepared to include concentrations from physiological orders of magnitude (discretized to a minimum, mean, and maximum) of each molecule found in the interstitial fluid and the blood, which were randomly drawn (49–53). In total, eight different baseline phantoms were prepared, representing individual skin types. Every liquid skin solution was then individually infused with ten randomized glucose concentrations chosen from an interval of 0-800 mg/dL (discretization steps of 10 mg/dL). Based on this process, in total 80 liquid skin phantoms were prepared for measurements, with precise chemical compositions as listed in **Supplementary Tab. 1**. In the liquid skin study, each sample was measured in 20 scans and subsequently averaged to calculate one OTS per sample. Instead of the autocalibration described above, a more simplistic calibration-based quantification approach was used to analyze the measured data, as the liquid skin samples are homogeneous solutions without layer-specific molecular contents. Each measurement was calibrated by subtracting a reference spectrum with a glucose concentration as close to 100 mg/dL from each measurement corresponding to the same skin type. After the calibration was carried out for each liquid skin type, a Partial Least Squares Regression model (5 components) combined with Recursive Feature Elimination (negative mean absolute error as decision criterion) with 5 features to select was trained for glucose estimation. For model testing, K-fold cross-validation with 5 splits was applied to determine the Mean Absolute Error (MAE), given as

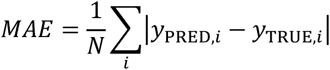

for evaluating the model accuracy between the predictions 𝑦_PRED,𝑖_ and ground truth values 𝑦_TRUE,𝑖_ for all 𝑁 = 80 datapoints. In addition to the MAE, the Mean Absolute Relative Difference (MARD), given as

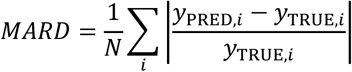

was reported. The computational processing pipeline for this study was implemented using Python, including scikit-learn.

### Multi-layer sample measurements

For the measurement of the PTFE-paper samples, 50, 100, and 200 µm thin PTFE layers were used from a circular aperture cell mount (6500S Buck Scientific) in combination with regular printing paper. 12 measurements with increasing camera trigger delay, i.e., 4 ms, 6 ms, 8 ms, 10 ms, 12 ms, 13.5 ms, 16 ms, 18 ms, 20 ms, 22 ms, 24 ms, and 26 ms after the start of each mid-IR irradiation period, were performed subsequently. For the biological multi-layer samples, mouse skin was cryosectioned to produce 40 µm and 60 µm thin slices of the skin sample. In the first experiment, the 60 µm skin sample was transferred onto the sample window and covered by a 10 µm thin PE foil to reduce drying of the sample and prevent water diffusion. 150 µL of water with glucose concentrations from 0 mg/dL to 10000 mg/dL in steps of 2500 mg/dL were subsequently pipetted onto the skin-foil-stack and scanned three times with a camera trigger delay of 13.5 ms and three times with a camera trigger delay of 19.5 ms. In a second experiment, the method was changed to capture multiple images, 4 ms, 13.5 ms, and 18.5 ms in one measurement. For this experiment, the 40 µm mouse skin was transferred onto the sample window and covered by a layer of 10 µm PE foil as described above. 150 µL of water with glucose concentrations of 0 mg/dL, 80 mg/dL, 110 mg/dL, 140 mg/dL, 170 mg/dL, 1000 mg/dL, and 10000 mg/dL were again pipetted onto the skin-foil-stack and measured subsequently with four scans per glucose concentration. Self-correction was performed by instead of determining 𝐹 by solving an optimization problem, by adjusting the magnitude of the absorption change in the skin layer based on the initial signal magnitude ratio between the 18.5 ms and the 4 ms measurement with

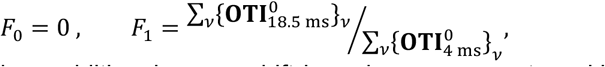

since the sample introduced no additional sensor drift based on movement or skin temperature over time.

### Human in vivo study

For the in vivo measurements in human volunteers, we received ethical approval from the TUM ethics commission filed in the German clinical trials register (DRKS00036371) in accordance with the declaration of Helsinki and ensured compliance with radiation protection law. All participants signed an informed consent before voluntary inclusion in the study. The study protocol includes measurements of the blood glucose and lactate levels in 20 healthy volunteers with our non-invasive optothermal system, as well as glucose additionally with a conventional minimally invasive enzymatic blood analyzer (Contour Next), and both glucose and lactate with a clinically grade blood analyzer (Biosen C-line EKF Diagnostics). In preparation for the measurements, the participants were told to fast for a minimum of 12 hours prior to the experiment. For the execution of the OGTT measurements, the volunteers were asked to position their wrist on the sample mirror of our optothermal spectroscopy system. Constant pressure was applied with the help of an inflatable cushion to hold the volunteers’ arms in place and to ensure stable experimental conditions. 25 min after the start of the experiment, the volunteers orally ingested a water solution of 300 mL with 75 g glucose (Gluko 75 Pulver, Medical Fox) to simulate food intake and the connected rise of blood glucose. During the experiment, the blood glucose level was measured invasively every 5 minutes over approximately 2 hours to collect a total of 25 reference glucose levels per OGTT. At the same time, four scans with our optothermal spectroscopy system were performed and averaged after every reference measurement. Throughout the measurements, the body temperature was monitored on the arm of the volunteers with a thermometer (54 II B, Fluke). The optothermal measurements were performed with three camera trigger delays of 4, 13.5, and 18.5 ms to capture data from different depths in the same scan. The data was subsequently processed in Python using the auto-calibration method as described above. The data from the 13.5 ms measurement, and therefore the data captured at the end of each mid-IR irradiation period, were corrected by the laser emission profile and analyzed in isolation as a comparison to the auto-calibrated data. For the analysis of spectra with comparable metabolite levels, 100 measurements with a reference blood glucose level close to 150 mg/dL were chosen from a pool of all measurements. The probe-beam instability was analyzed by calculating the geometric center of the full-width half-maximum beam area captured at the beginning of each mid-IR irradiation cycle.

## Data Availability

The data that support the findings of this study are available from the corresponding authors upon reasonable request.

## Acknowledgements

We gratefully acknowledge Leonard Reidiess for his contribution to initial system implementation and characterization, and Aleksandra Kolbasnikova for conducting phantom experiments. Furthermore, we thank Dr. Ute Chambers and Dr. Ulrich Stahl for their efforts in obtaining ethical study approval. This project has received funding from the Helmholtz Munich Innovation & Translation Call (OPTO-G); from the TranslaTUM Seed Fund (Project Deep-OT); the European Research Council (ERC) under the European Union’s Horizon Europe research and innovation programme under grant agreement No 101041936 (EchoLux); and from the European Union’s Horizon Europe research and innovation programme under grant agreement No 101058111 (GLUMON).

## Author contributions

Conceptualization: CB, BP, SH, MAP

Formal analysis: CB, BP

Funding acquisition: MAP

Investigation: CB, BP, IH, SH

Methodology: CB, BP

Project administration: CB, MAP

Software: CB, BP, IH

Resources: MAP

Supervision: DJ, MAP

Validation: CB, BP, JQ

Visualization: CB, BP

Writing–original draft: CB, BP, MAP

Writing–review & editing: CB, BP, JQ, IH, SH, DJ, MAP

## Supplementary figures

**Supplementary Fig. 1.**
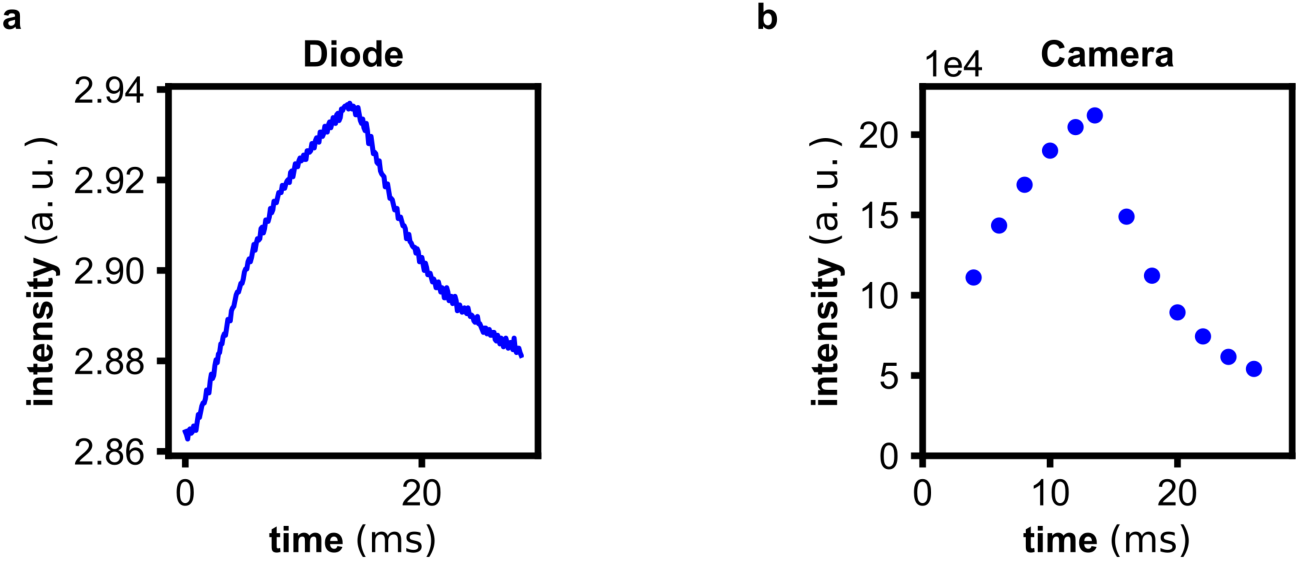
Heat-up cool-down curve captured with a single point detector and a camera. A drop of water was placed on the sample window and irradiated by a mid-IR laser for 13.5 ms, leading to a perturbation of the probe beam, which was quantified using a single point detector (a) and a camera (b).

**Supplementary Fig. 2.**
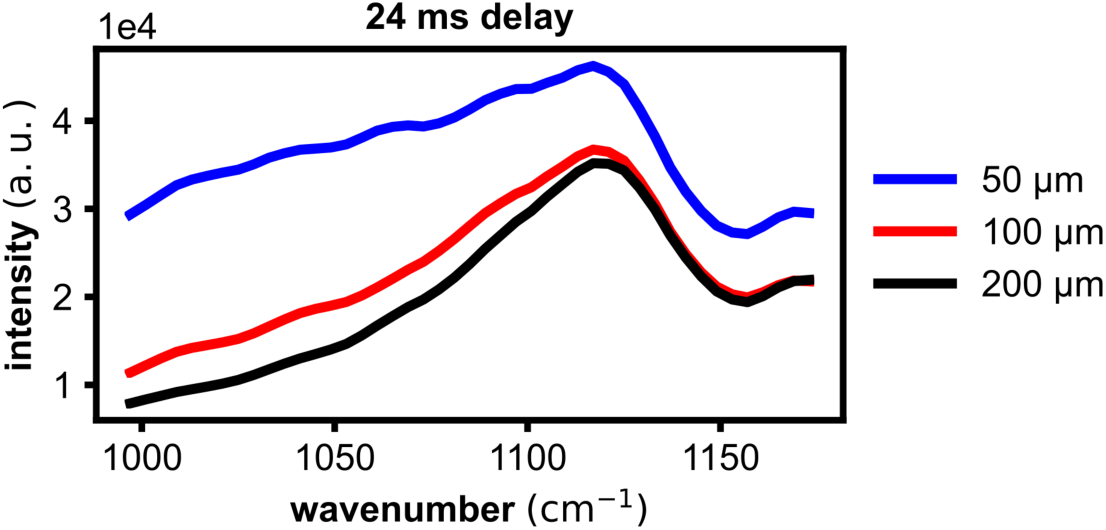
System response for three PTFE-paper stacks with increasing PTFE thickness. The system responses were captured using an acquisition delay of 24 ms.

**Supplementary Fig. 3.**
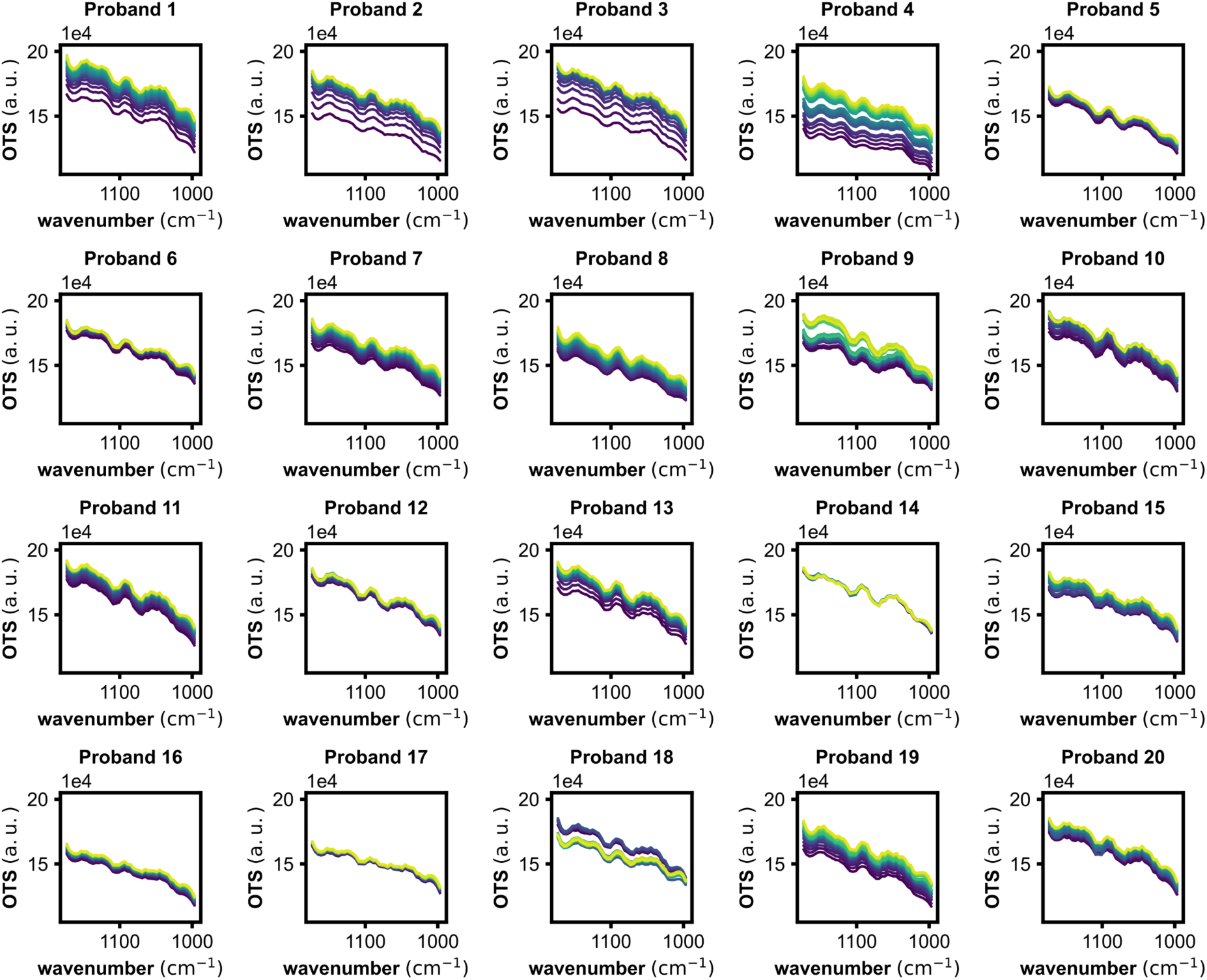
Non-corrected spectra of all volunteer measurements. The spectra were acquired based on an acquisition delay of 13.5 ms, which corresponds to the mid-infrared irradiation cycle.

**Supplementary Fig. 4.**
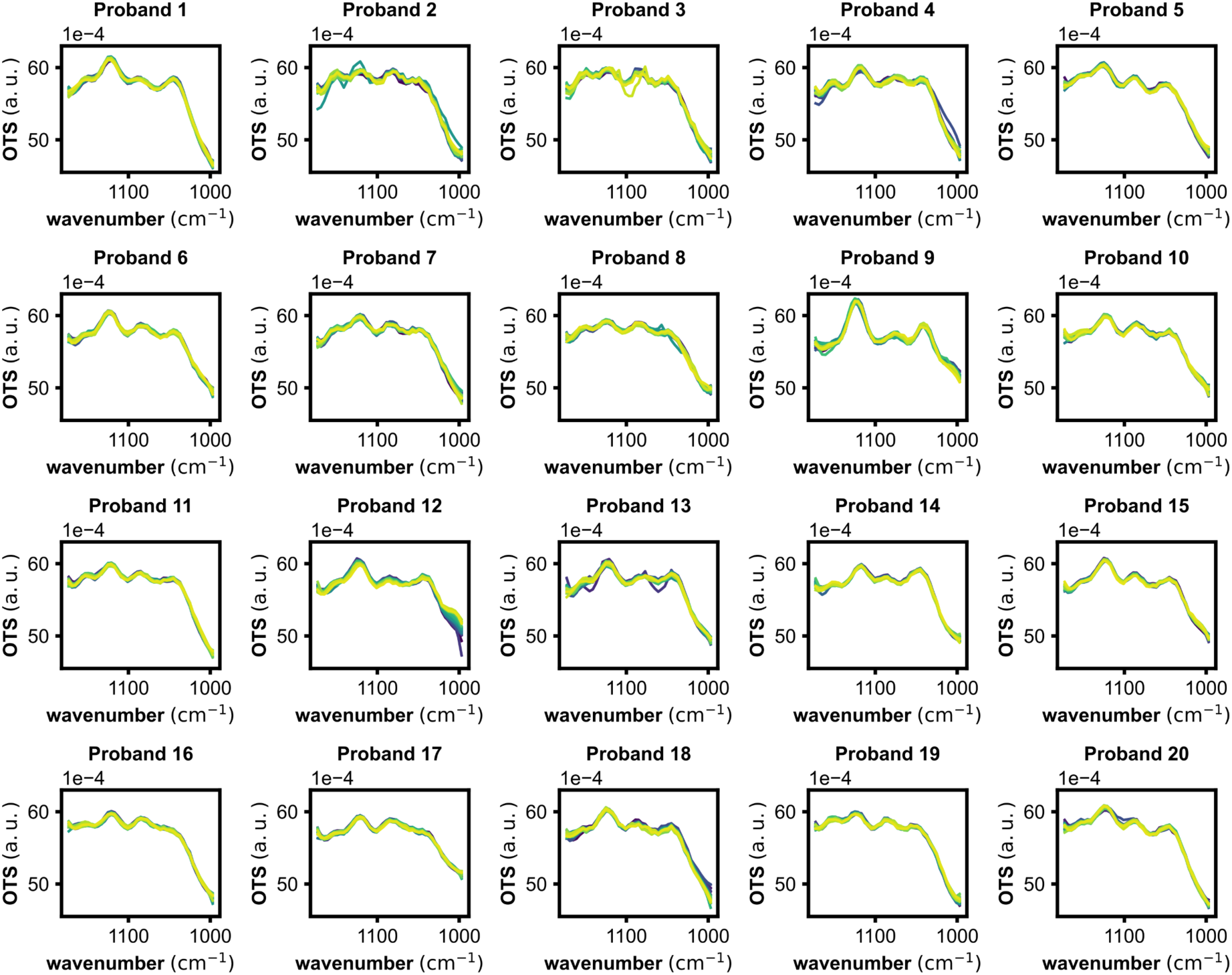
**Autocorrected spectra of all volunteer measurements.**

**Supplementary Fig. 5.**
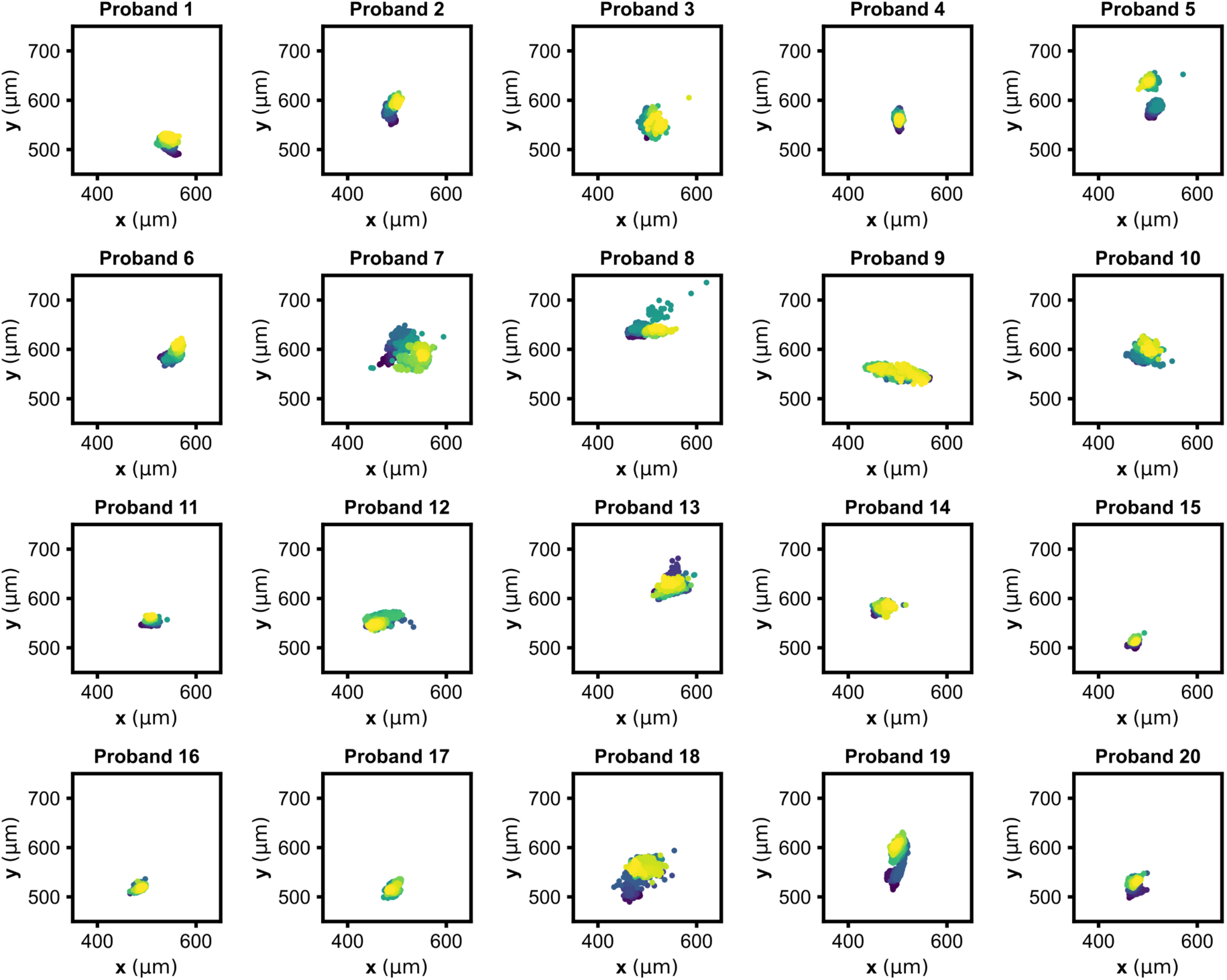
**Probe beam wandering over the course of each volunteer measurement.**

**Supplementary Tab. 1.** Liquid skin phantom.

| *Skin type* | *Albumin [mg/dL]* | *Lactate [mg/dL]* | *Intralipid [mg/dL]* | *Glucose superpositions [mg/dL]* |  |  |  |  |  |  |  |  |  |
| --- | --- | --- | --- | --- | --- | --- | --- | --- | --- | --- | --- | --- | --- |
|  |  |  |  | 1 | 2 | 3 | 4 | 5 | 6 | 7 | 8 | 9 | 10 |
| 1 | 1860 | 105 | 3333 | 10 | 160 | 250 | 300 | 300 | 450 | 490 | 560 | 610 | 700 |
| 2 | 1860 | 200 | 3333 | 120 | 130 | 150 | 180 | 190 | 370 | 430 | 450 | 540 | 760 |
| 3 | 5600 | 200 | 5167 | 20 | 300 | 340 | 350 | 390 | 400 | 450 | 460 | 510 | 780 |
| 4 | 3730 | 200 | 7000 | 20 | 50 | 130 | 250 | 350 | 370 | 500 | 520 | 570 | 580 |
| 5 | 1860 | 200 | 3333 | 150 | 230 | 530 | 530 | 570 | 620 | 620 | 730 | 740 | 740 |
| 6 | 3730 | 200 | 7000 | 130 | 200 | 230 | 370 | 470 | 480 | 520 | 530 | 620 | 700 |
| 7 | 1860 | 10 | 7000 | 40 | 50 | 90 | 90 | 150 | 260 | 400 | 570 | 610 | 790 |
| 8 | 5600 | 200 | 5167 | 40 | 70 | 90 | 100 | 270 | 290 | 290 | 300 | 410 | 520 |

